# Adolescent engagement and sentiment toward reproductive health videos on Chinese social media: A cross-sectional content analysis

**DOI:** 10.1101/2025.06.18.25329831

**Authors:** Xuening Zhang, Zixuan Zhou, Jianhao Ma, Yu Chen, Yifan Xu, Zhenxu Cui, Jinpeng Li, Jieer Qiu, Qinglong Zhang, Ruochen Huang

## Abstract

Adolescents increasingly rely on social media for reproductive health information, yet the scope, quality, and emotional reception of such content remain poorly characterized in China. We analysed 743 Bilibili videos published between September 2016 and June 2024, encompassing 486.8 million views and approximately 760,000 comments and danmaku, using a 29-keyword query. Pearson correlation, descriptive statistics, and large-language-model (Qwen2.5-32B) sentiment classification were applied to engagement metrics and user-generated text; uploader medical credentials were manually verified. Views correlated strongly with likes (*r* = 0.70), collections (*r* = 0.74), and shares (*r* = 0.74). Only 17% of uploaders had confirmed medical training, with no medically qualified creators identified for novel-contraceptive or spontaneous-abortion content. Niche topics such as sterilization surgery (3.0% of videos) and fertility-awareness methods (2.3%) attracted disproportionately high engagement. Negative sentiment predominated for induced abortion (52.2%) and novel contraceptives (52.1%), whereas coping with spontaneous abortion elicited the highest positive response (24.2%). Negative sentiment correlated moderately with comment volume (*r* = 0.32) but inversely with likes (*r* = *−*0.38). Adolescent engagement on Bilibili centers on basic contraception, yet contentious or under-covered topics drive intense, emotionally polarised interactions largely without professional oversight. Collaboration among healthcare providers, educators, and influential creators is needed to ensure accurate, empathetic content that meets adolescents’ unmet informational needs.

## Introduction

### Background

Adolescent reproductive health is fundamental to the physical and psychological well-being of young individuals. It addresses a broad spectrum of needs, including access to contraception, safe abortion services, and the prevention of sexually transmitted diseases (STDs) [1]. Despite notable progress in women’s health over recent decades, traditional healthcare systems frequently fall short in addressing adolescents’ diverse reproductive health needs, particularly in settings such as China [2]. The limited availability and sensitivity surrounding these topics in conventional educational environments often results in substantial gaps in adolescents’ understanding and preparedness to manage reproductive health challenges effectively [3].

The rapid expansion of the internet and social media has fundamentally altered how adolescents seek health-related information, especially regarding sensitive and stigmatized topics. Digital platforms have emerged as indispensable resources for reproductive health education, providing anonymity, convenience, and accessibility that resonate strongly with younger populations [4–6]. Previous studies demonstrate the effectiveness of digital interventions, including online educational programs and mobile health applications, in enhancing adolescents’ sexual health knowledge and influencing positive behavioral changes [7–10]. By offering a scalable and discrete alternative, online platforms can potentially overcome many limitations associated with traditional reproductive health service delivery.

### Rationale and study context

In China, Bilibili, a video-sharing platform particularly popular among Generation Z, has become a central medium for young people seeking reproductive health information. As of 2024, Bilibili boasts approximately 341 million monthly active users, more than 80% of whom belong to the younger demographic group aged 24 years or below [4]. This unique user profile underscores Bilibili’s potential as a powerful channel for adolescent health communication, highlighting the need for understanding the nature and effectiveness of reproductive health content available on this platform.

To explore adolescents’ reproductive health concerns and needs, this study leverages web text mining to analyze user-generated content on Bilibili. Web text mining has proven valuable in public health research for extracting meaningful insights from social media data, such as identifying health-related concerns and information gaps [11–13]. By examining comments and discussions linked to reproductive health videos on Bilibili, this research seeks to uncover data-driven insights that can inform targeted policy development and service enhancements for adolescent reproductive health.

### Objective

Given the critical role of digital platforms in adolescent health education, it is vital to ensure the reliability and comprehensiveness of information disseminated online. However, concerns persist regarding the credibility and quality of health-related content available on social media, often produced without medical oversight [14, 15]. By examining the patterns of user engagement, content coverage, uploader credibility, and viewer sentiment on Bilibili, this study aims to identify significant informational needs, content gaps, and emotional influences shaping adolescent reproductive health discourse online. These insights could inform targeted strategies to enhance the quality and impact of digital reproductive health interventions, ensuring that adolescents receive accurate, empathetic, and comprehensive information.

## Methods

### Data source and collection

A total of 743 videos focusing on reproductive health topics were retrospectively collected from Bilibili. These videos were uploaded between September 2016 and June 2024, with each having accumulated at least 100,000 views to ensure historical breadth and capture evolving public health trends over time. The video retrieval process relied on a predefined set of search keywords spanning two primary categories: contraception-related terms (e.g., “安全套” [Condom], “ 避孕药” [Oral Contraceptives], “节育器” [Intrauterine Device]) and abortion-related terms (e.g., “人工流产” [Induced Abortion], “自然流产” [Spontaneous Abortion]). Additional keywords related to general reproductive health education were also incorporated. These keywords were systematically applied to video titles and user-generated tags to identify candidate content; the comprehensive keyword framework is detailed in S1 Table.

Video metadata and user-generated textual data (including both standard comments and real-time danmaku) were retrieved for each video.A distributed web-crawling framework was developed to systematically retrieve the Bilibili video data and transfer it to a relational MySQL database for persistent storage. For each included video (*n* = 743), two distinct layers of information were extracted:

- **Structural metadata:** Video title, upload timestamp, uploader unique ID, uploader follower count, and a comprehensive suite of user engagement metrics, including views (total view count), likes (number of thumbs-up), shares (forwarding count), collections (bookmarking count), coins (virtual currency rewarded to the uploader), comments (total text comments), and danmaku (real-time synchronized overlay comments).
- **User-generated text:** The full-text programmatic export of all comments and danmaku streams associated with the respective videos.

After collection, two researchers independently screened every clip to confirm topical relevance and removed items unrelated to reproductive health as shown in Figure 1. The final dataset comprises unique uploaders (*n* = 525) and distinct viewers (*n* = 218, 000), together logging views (*n* = 486.83 million), likes (*n* = 12.31 million), collections (*n* = 3.35 million), coins (*n* = 1.76 million), and shares (*n* = 3.19 million). It also retains comments (*n* = 328, 256) and danmaku (*n* = 432, 116) alongside complete metadata and textual artefacts for all downstream analyses.

**Fig 1.** Search strategy and video screening procedure.

### Classification of videos

Video uploaders were classified into two groups: (1) medical-background uploaders and (2) non-medical-background uploaders. The identities of medical-background and non-medical-background uploaders were distinguished by systematically reviewing each creator’s self-introductions and related videos: we extracted any self-reported clinical titles, specialties, institutional affiliations, and other credential-relevant information displayed within the videos, classifying those with verifiable medical details as medical-background uploaders and those without as non-medical-background uploaders.

Video content was classified into 13 categories: (1) Condom, (2) Female Condom, (3) Device-based Contraception, (4) Coitus Interruptus, (5) Contraception Education, (6) Oral Contraceptives, (7) Sterilization Surgery, (8) Fertility Awareness-Based Methods, (9) Novel Contraceptive Methods, (10) Contraceptive Failure, (11) Induced Abortion, (12) Spontaneous Abortion, and (13) Reproductive Health Education.

The detailed criteria and definitions for each publisher type and content category are provided in Table 1. All videos were independently reviewed and classified by two researchers, and any disagreements were resolved through discussion or consultation with a third reviewer.

**Table 1.** Classification of video uploaders and content.

| Category | Description |
| --- | --- |
| *Video Publishers* |  |
| Medical-background uploaders | Content creators whose professional identity can be verified as belonging to licensed healthcare occupations, such as physicians, nurses, midwives, reproductive endocrinologists, or other allied health practitioners, through self introductions, institutional affiliations, or medically themed portfolios. Their formal clinical training (for example, completion of accredited medical programs and possession of practice certificates) generally confers higher credibility and accuracy to reproductive health information shared on social media platforms. |
| Non-medical-background uploaders | Individuals who publish reproductive health content without verifiable clinical credentials. This group includes science communicators, health educators, teachers, popular science vloggers, and other enthusiasts lacking formal medical licensure or training. While they may contribute valuable educational perspectives, the absence of certified healthcare expertise can limit the reliability of the information they provide. |
| *Video Content* |  |
| Condom | Including types and brands of condoms, correct usage techniques, common misconceptions, purchasing tips, disease-prevention efficacy, contraceptive knowledge, and related anecdotes or social phenomena. |
| Female Condom | Including appearance and structure of female condoms, insertion and removal procedures, real-world experience, and relevant popular-science information. |
| Device-based Contraception | Including principles, user experiences, advantages and disadvantages, common misconceptions, and social discussions surrounding device-based methods such as intrauterine devices. |
| Coitus Interruptus | Including contraceptive principles, pregnancy risks, safety considerations for both partners, typical effectiveness, and widespread misconceptions about withdrawal. |
| Contraception Education | Including overviews of various contraceptive methods, instructions for correct use, the importance of protection, common misconceptions, and foundational sex-education knowledge. |
| Oral Contraceptives | Including pharmacologic principles, user experiences, side effects, common misconceptions, and recent advances in hormone-based methods such as male pills, implants, and injectable contraceptives. |
| Sterilization Surgery | Including procedures, mechanisms, advantages, disadvantages, and social discourse surrounding male and female sterilization surgeries. |
| Fertility Awareness-Based Methods (FABMs) | Including principles of safe-period tracking, pregnancy risks, limitations, popular-science explanations, and frequent misunderstandings of fertility awareness methods. |
| Novel Contraceptive Methods | Including principles, user experiences, comparative analyses, and notable anecdotes related to emerging contraceptive tools and unconventional approaches. |
| Contraceptive Failure | Including real cases of unintended pregnancy, root-cause analyses, prevalent misconceptions, and recommended responses to contraceptive failure. |
| Induced Abortion | Including medical and surgical procedures, associated risks and physical impacts, relevant social phenomena, and the importance of contraception to prevent unintended pregnancies. |
| Spontaneous Abortion | Including personal accounts, emotional and health impacts, and medical contexts of spontaneous abortion due to fetal demise, accidents, or maternal illness. |
| Reproductive Health Education | Including comprehensive information on sex and reproduction such as contraception, ejaculation, pregnancy, abortion, gynecological health, fertility, stretch marks, and sexual skills, promoting scientific attitudes and healthy concepts. |

### Data preprocessing

To ensure data quality and analytical reliability, several preprocessing steps were applied to the collected video metadata, comments, and danmaku. These steps addressed issues such as redundancy, irrelevance, and linguistic complexity of Chinese text.

1. *Duplicate removal* : Duplicate entries were identified and eliminated using unique identifiers. For videos, duplicates were detected via video IDs; for comments and danmaku, comment IDs and danmaku IDs were used.
2. *Irrelevant Data Filtering*: Comments and danmaku unrelated to reproductive health were filtered out to maintain focus. A large language model (LLM) assisted in automatically tagging and removing irrelevant content, supplemented by manual sampling to validate the filtering process.
3. *Text Processing for Chinese Language*: Given that Chinese text lacks spaces between words, word segmentation was performed using the Jieba library, a widely adopted tool for Chinese natural language processing. This step enabled accurate tokenization of comments and danmaku. Colloquial expressions, slang, and emojis prevalent in danmaku were addressed by either removing them or converting emojis to textual descriptions (e.g., “smile” for a smiley face) to preserve contextual meaning.

These preprocessing steps resulted in a cleaned, structured dataset suitable for in-depth analysis, minimizing noise and enhancing the focus on reproductive health-related insights.

### Analysis methods

Multiple analytical techniques were employed to explore the data and address the study’s objectives of understanding adolescent reproductive health concerns and engagement patterns on Bilibili. These methods are detailed below.

#### Descriptive and correlation analysis

To assess user engagement, a descriptive analysis was conducted on key video metrics: view, likes, shares, collections, coins, comments, and danmaku. This provided a quantitative overview of interaction patterns across the 743 videos.

A correlation analysis was conducted to explore relationships among the engagement metrics. Pearson’s correlation coefficient was calculated to construct a correlation matrix, quantifying linear relationships between pairs of variables (e.g., view vs. likes, likes vs. collections). The coefficient ranges from *−*1 (strong negative correlation) to 1 (strong positive correlation), with 0 indicating no correlation. For visualization, a scatter plot matrix was generated, plotting each metric against others to reveal trends and outliers. This analysis illuminated engagement dynamics, such as whether videos with higher views consistently received more likes or comments.

#### Sentiment analysis

To explore the emotional tone within user discussions about reproductive health, we conducted a sentiment analysis on comments and danmaku [16]. For this purpose, we employed the LLM Qwen2.5-32B to classify each text entry into one of three sentiment categories: positive, negative, or neutral [17].

The Qwen2.5-32B model was chosen due to its strong capability in handling Chinese text, especially the informal and colloquial expressions often found in online conversations [18]. This made it an ideal tool for evaluating user attitudes toward topics like contraception and abortion. By classifying the sentiments, we gained a clear picture of the prevailing trends, shedding light on users’ general experiences and concerns related to these reproductive health themes [19].

Together, these analytical approaches provide a robust framework for analyzing Bilibili data, combining quantitative engagement metrics, thematic exploration, and emotional insights to comprehensively address adolescent reproductive health needs.

### Ethical considerations

The data analyzed in this research included only openly accessible content, such as video metadata, comments, and danmaku. Importantly, no personal identifying information was collected beyond user aliases, ensuring that the privacy of individuals remained fully protected throughout the study. To prioritize user privacy, the research team took deliberate steps to avoid publishing or sharing raw user comments or danmaku in any form. Instead, the findings presented in this study are derived from aggregated and anonymized data. This approach guarantees that no individual user can be identified from the results, further safeguarding privacy. Since the research relied exclusively on publicly available and anonymized data, a formal ethics review was deemed unnecessary. However, the research team remained dedicated to responsible data practices. This included securely storing the dataset and following best practices for data protection to mitigate any potential risks.

## Results

### Relationship between video features and user engagement

To examine the relationships between video features and user engagement behaviors in online health videos, we performed a Pearson correlation analysis using a data matrix comprising key variables. The analysis aimed to quantify the strength and direction of associations between these variables (Figure 2).

**Fig 2.** Correlation matrix of video features.

The results revealed several notable patterns. Views exhibited strong positive correlations with multiple engagement metrics, including likes (*r* = 0.70), collections (*r* = 0.74), and shares (*r* = 0.74). This suggests that videos with higher view counts are more likely to receive likes, be saved by users, and be shared with others. Similarly, likes showed strong positive correlations with collections (*r* = 0.86) and shares (*r* = 0.82), indicating that videos receiving more likes are also frequently saved and shared.

A particularly strong correlation was observed between collections and shares (*r* = 0.90), highlighting a robust association between users saving a video and sharing it. Additionally, coins were moderately to strongly correlated with likes (*r* = 0.74), collections (*r* = 0.65), and danmaku (*r* = 0.68), suggesting that videos which receive more likes or are saved more often also tend to receive more financial support and have higher levels of real-time commenting activity.

Comments and danmaku demonstrated a moderate positive correlation (*r* = 0.63), indicating that videos with higher comment counts tend to have increased danmaku. However, the relationship between views and danmaku was weaker (*r* = 0.29), indicating that viewership does not effectively drive real-time commenting activity.

In contrast, uploader followers showed only weak to moderate correlations with most engagement metrics, with the strongest association being with danmaku (*r* = 0.46). This suggests that the uploader’s follower count has limited influence on engagement behaviors such as liking, sharing, or commenting on specific videos.

To further elucidate these relationships, we employed a scatterplot matrix to visually represent the pairwise associations among the selected engagement metrics (Figure 3). Each subplot in the matrix depicted the relationship between two variables, with a red regression line indicating the trend and a pink confidence interval reflecting the precision of the estimate. The scatterplot matrix confirmed the strong positive relationships identified in the correlation analysis, particularly between views, likes, collections, and shares, where data points clustered tightly along upward-trending regression lines in the low to moderate range.

**Fig 3.** Scatter-plot matrix of video features.

However, in high-engagement regions (e.g., views exceeding 10 million), data points became sparser, and confidence intervals widened, revealing increased variability and potential outliers. Additionally, the weaker influence of uploader followers was visually evident through flatter regression lines and wider confidence intervals, emphasizing that content quality outweighs uploader popularity in driving engagement.

### Adolescents’ attention to different reproductive health topics

Following the analysis of engagement metrics, we now explore their variation across reproductive health topics on Bilibili. In this study, we divided the videos into 13 categories, as shown in Table 2, and represented them in the form of tags based on keyword classification and content analysis. By examining topic distribution, trends, and uploader influence, we aim to pinpoint content that captivates adolescents. These insights are key to understanding teen interests and refining educational strategies for effective outreach.

**Table 2.** Distribution of videos across tag categories.

| Tag | Ratio (%) | *n* |
| --- | --- | --- |
| Condom | 37.8 | 280 |
| Reproductive Health Education | 20.1 | 149 |
| Induced Abortion | 8.7 | 64 |
| Contraceptive Education | 7.9 | 58 |
| Oral Contraceptives | 6.9 | 51 |
| Contraceptive Failure | 5.4 | 40 |
| Device-based Contraception | 3.7 | 27 |
| Sterilization Surgery | 3.0 | 22 |
| Fertility Awareness-Based Methods (FABMs) | 2.3 | 17 |
| Coitus Interruptus | 1.8 | 13 |
| Female Condom | 1.1 | 8 |
| Spontaneous Abortion | 1.0 | 7 |
| Novel Contraceptive Methods | 0.9 | 6 |

The distribution of reproductive health videos on Bilibili reveals a clear concentration on specific topics. As shown in Table 2, videos tagged with “Condoms” and “Reproductive Health Education” dominate, collectively accounting for 57.9%(430*/*743) of the total sample. This suggests that adolescents’ exposure to reproductive health content is heavily skewed toward basic contraceptive methods and general education, reflecting uploader priorities.

In contrast, other critical topics such as “Female Condom,” “Spontaneous Abortion,” and “Novel Contraceptive Methods” constitute a significantly smaller proportion of the content. This uneven distribution indicates potential gaps in the availability of information on more specialized or sensitive reproductive health issues, which may limit adolescents’ access to comprehensive knowledge in these areas.

Temporal trends in video production, illustrated in Figure 4, show a marked increase in the number of reproductive health videos from 2016 to 2024, although the apparent decline in 2024 occurs because our dataset includes only the first half of that year. This upward trend suggests growing interest in reproductive health topics, possibly reflecting increased awareness or demand among adolescent viewers during this period.

**Fig 4.** Temporal trend in the number of tagged videos.

However, growth in other topics, such as “FABMs,” “Sterilization Surgery,” and “Coitus Interruptus,” remained limited over the same timeframe. This disparity highlights an imbalance in content coverage, where the proliferation of videos on popular topics outpaces that of less-covered but equally important subjects. This trend may reflect insufficient focus by uploaders on these areas, potentially exacerbating information gaps.

User engagement metrics, specifically like rate (likes per view), comment rate (comments per view), and share rate (shares per view), provide valuable insights into how various reproductive health topics engage viewers. Table 3 summarizes these metrics across tags, revealing distinct patterns of interaction.

**Table 3.** User engagement metrics by content tag.

| Tag | Like Rate (%) | Comment Rate (%) | Share Rate (%) |
| --- | --- | --- | --- |
| Induced Abortion | 4.35 | 0.42 | 0.70 |
| Sterilization Surgery | 3.96 | 0.20 | 0.58 |
| Oral Contraceptives | 3.56 | 0.35 | 1.11 |
| FABMs | 2.94 | 0.16 | 0.70 |
| Contraceptive Education | 2.87 | 0.20 | 0.64 |
| Contraceptive Failure | 2.64 | 0.19 | 0.28 |
| Female Condom | 2.60 | 0.16 | 0.46 |
| Reproductive Health Education | 2.44 | 0.20 | 0.61 |
| Condom | 2.41 | 0.15 | 0.60 |
| Spontaneous Abortion | 2.40 | 0.20 | 0.27 |
| Device-based Contraception | 2.38 | 0.22 | 0.47 |
| Coitus Interruptus | 1.26 | 0.26 | 0.75 |
| Novel Contraceptive Methods | 1.20 | 0.23 | 0.39 |

The data indicates that topics such as “Induced Abortion,” “Sterilization Surgery,” and “Oral Contraceptives” consistently elicit heightened engagement across all three metrics. For instance, “Induced Abortion” records a like rate of 4.35%, a comment rate of 0.42%, and a share rate of 0.70%, positioning it as a standout topic in terms of viewer resonance, discussion potential, and perceived value. Similarly, “Sterilization Surgery” achieves a like rate of 3.96%, paired with a moderate share rate of 0.58%, while “Oral Contraceptives” exhibits a like rate of 3.56%, a comment rate of 0.35%, and the highest share rate of 1.11%. These figures suggest that viewers not only approve of such content but also find it compelling enough to discuss and share, likely due to its emotional complexity, practical relevance, or informational depth. Other specialized topics, such as “FABMs” (like rate: 2.94%, share rate: 0.70%) and “Coitus Interruptus” (like rate: 1.26%, comment rate: 0.26%, share rate: 0.75%), further reinforce this trend, showing notable engagement despite their lower prevalence on the platform.

In contrast, more familiar and widely covered topics like “Condoms” and “Reproductive Health Education” exhibit consistently lower engagement metrics. “Condoms” records a like rate of 2.41%, a comment rate of 0.15%, and a share rate of 0.60%, while “Reproductive Health Education” shows a like rate of 2.44%, a comment rate of 0.20%, and a share rate of 0.61%. These subdued responses suggest that their broad coverage may lead to saturation, reducing their novelty and capacity to provoke strong reactions or discussions among viewers. This disparity underscores a key finding: while prevalent topics dominate in volume, they do not necessarily align with the intensity of viewer interest or interaction observed in less-represented subjects.

Given that the credibility and accuracy of reproductive health content heavily depend on the professional knowledge of uploaders, analyzing their backgrounds across various reproductive health topics is crucial (Table 4). This analysis reveals substantial variability in medical expertise among uploaders, influencing both content reliability and viewer engagement. Despite the critical importance of professional credibility, our manual confirmation of the medical backgrounds of 525 uploaders through their self-introductions and related videos reveals that the overall proportion of uploaders with medical backgrounds in adolescent reproductive health education on Bilibili remains relatively low (17%, 89/525). This finding raises significant concerns regarding the quality and reliability of educational content provided on the platform, underscoring the necessity for greater involvement of medically qualified contributors to ensure information accuracy and enhance public health outcomes.

**Table 4.**
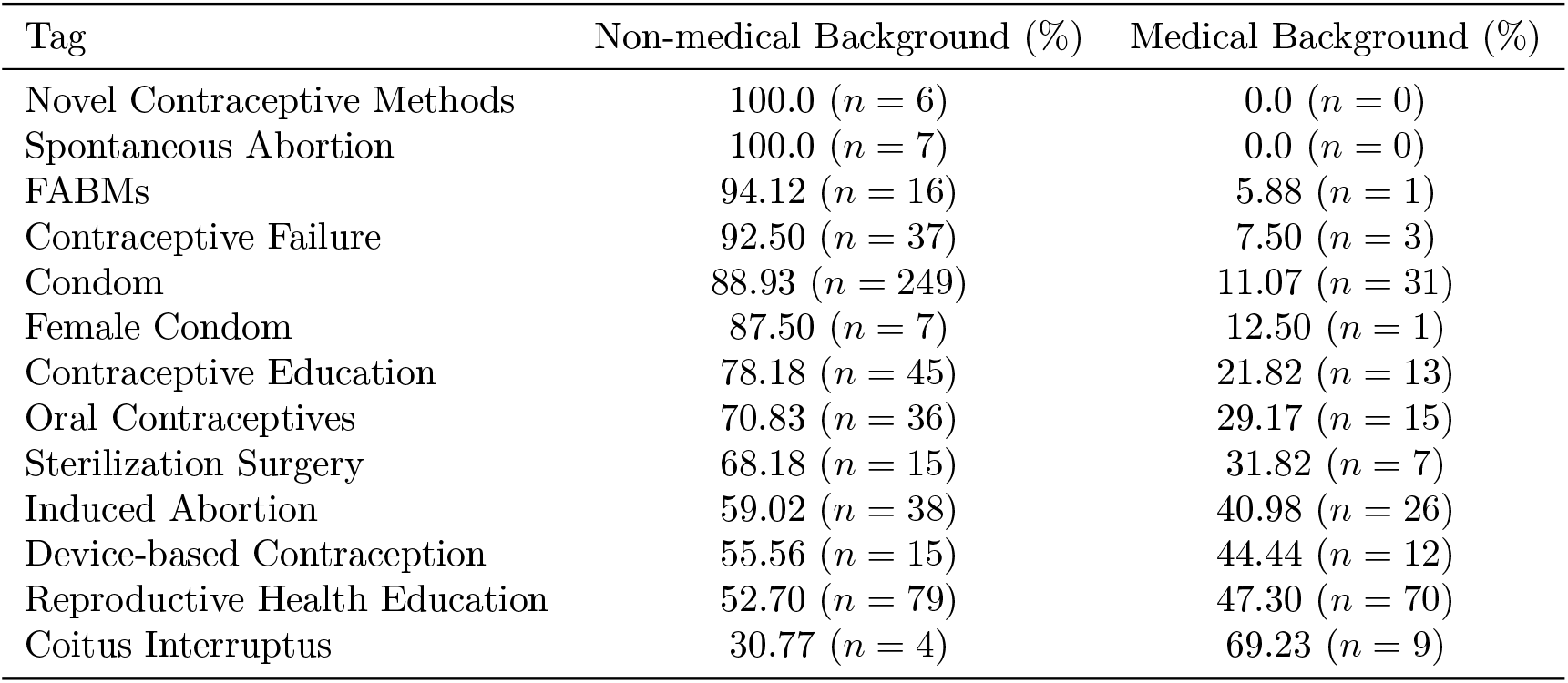
Distribution of medical background among uploaders across tag categories.

| Tag | Non-medical Background (%) | Medical Background (%) |
| --- | --- | --- |
| Novel Contraceptive Methods | 100.0 ($n = 6$) | 0.0 ($n = 0$) |
| Spontaneous Abortion | 100.0 ($n = 7$) | 0.0 ($n = 0$) |
| FABMs | 94.12 ($n = 16$) | 5.88 ($n = 1$) |
| Contraceptive Failure | 92.50 ($n = 37$) | 7.50 ($n = 3$) |
| Condom | 88.93 ($n = 249$) | 11.07 ($n = 31$) |
| Female Condom | 87.50 ($n = 7$) | 12.50 ($n = 1$) |
| Contraceptive Education | 78.18 ($n = 45$) | 21.82 ($n = 13$) |
| Oral Contraceptives | 70.83 ($n = 36$) | 29.17 ($n = 15$) |
| Sterilization Surgery | 68.18 ($n = 15$) | 31.82 ($n = 7$) |
| Induced Abortion | 59.02 ($n = 38$) | 40.98 ($n = 26$) |
| Device-based Contraception | 55.56 ($n = 15$) | 44.44 ($n = 12$) |
| Reproductive Health Education | 52.70 ($n = 79$) | 47.30 ($n = 70$) |
| Coitus Interruptus | 30.77 ($n = 4$) | 69.23 ($n = 9$) |

The analysis reveals substantial variability in medical expertise among uploaders across various topics. “Coitus Interruptus” shows the highest medical representation at 69.23%(9/13), indicating strong professional engagement. Other topics like “Reproductive Health Education” and “Device-based Contraception” also have considerable medical involvement, with proportions of 47.30%(70/149) and 44.44%(12/27), respectively.

Furthermore, “Induced Abortion” and “Sterilization Surgery” reflect significant professional presence, with medical uploaders accounting for 40.98%(26/64) and 31.82%(7/22).

In stark contrast, topics such as “Condoms” exhibit minimal medical involvement, with only 11.07%(31/280) of uploaders having medical backgrounds, suggesting a predominance of non-expert uploaders (249/280, 88.93%). More alarmingly, content categories including “Novel Contraceptive Methods” and “Spontaneous Abortion” entirely lack contributions from medically trained professionals, raising serious questions about content reliability and safety.

### User sentiment responses to reproductive health content

The analysis of user sentiment responses, classified through a LLM, revealed distinct emotional reactions to various reproductive health topics (Table 5).

**Table 5.** Sentiment analysis across different tags.

| Tag | Negative (%) | Neutral (%) | Positive (%) |
| --- | --- | --- | --- |
| Induced Abortion | 52.22 ($n = 58,347$) | 30.87 ($n = 34,492$) | 16.91 ($n = 18,894$) |
| Novel Contraceptive Methods | 54.38 ($n = 2,530$) | 33.69 ($n = 1,568$) | 11.93 ($n = 556$) |
| Sterilization Surgery | 50.80 ($n = 5,960$) | 33.10 ($n = 3,883$) | 16.10 ($n = 1,889$) |
| Device-based Contraception | 50.70 ($n = 18,606$) | 35.17 ($n = 12,907$) | 14.13 ($n = 5,185$) |
| Coitus Interruptus | 48.68 ($n = 4,943$) | 41.36 ($n = 4,200$) | 9.96 ($n = 1,011$) |
| Contraceptive Failure | 48.13 ($n = 36,743$) | 29.85 ($n = 22,620$) | 22.02 ($n = 16,686$) |
| Oral Contraceptives | 47.16 ($n = 25,244$) | 37.41 ($n = 20,025$) | 15.42 ($n = 8,254$) |
| FABMs | 46.51 ($n = 4,448$) | 37.40 ($n = 3,577$) | 16.09 ($n = 1,539$) |
| Reproductive Health Education | 43.67 ($n = 49,130$) | 37.53 ($n = 42,222$) | 18.80 ($n = 21,150$) |
| Condom | 42.97 ($n = 107,474$) | 41.74 ($n = 104,398$) | 15.29 ($n = 38,242$) |
| Female Condom | 41.42 ($n = 2,291$) | 42.79 ($n = 2,367$) | 15.79 ($n = 873$) |
| Contraceptive Education | 40.14 ($n = 26,659$) | 39.37 ($n = 26,147$) | 20.50 ($n = 13,615$) |
| Spontaneous Abortion | 38.38 ($n = 4,504$) | 37.39 ($n = 4,388$) | 24.23 ($n = 2,843$) |

Topics such as “Induced Abortion” and “Novel Contraceptive Methods” elicited predominantly negative sentiments, accounting for 52.22%(58347/111733) and 54.38%(2530/4654), respectively. These elevated negative responses may reflect societal controversies or apprehensions related to ethical considerations, acceptance of emerging technologies, or concerns regarding side effects and irreversible outcomes associated with these methods. Similar trends were observed for “Sterilization Surgery” and “Device-based Contraception,” which also showed high negative sentiment percentages (5960/11732,50.80% and 18606/36698,50.70%, respectively).

In contrast, topics like “Spontaneous Abortion” exhibited the highest proportion of positive sentiments (2843/11735, 24.23%), indicating users’ empathy, support, and understanding towards recovery experiences. Additionally, “Reproductive Health Education” content garnered considerable positive sentiment (21150/112502,18.80%), highlighting the appreciation for educational efforts in promoting reproductive health awareness.

The study further explored how different sentiment categories correlate with user engagement behaviors (Figure 5). Negative sentiment displayed a moderate positive correlation with comments (*r* = 0.32), indicating that contentious or controversial content typically fosters more extensive discussions. Conversely, negative sentiment correlated negatively with supportive user interactions such as likes (*r* = *−*0.38) and coins (*r* = *−*0.51), suggesting that negative content might inhibit positive engagement behaviors.

**Fig 5.** Correlation heatmap of sentiment percentages and video metrics.

Neutral sentiment showed moderate positive correlations with views (*r* = 0.32), implying such content attracts attention but may not stimulate extensive interactions or deep discussions, as evidenced by its negative correlation with comments (*r* = *−*0.32). However, neutral content demonstrated a relatively strong positive correlation with shares (*r* = 0.40), signifying that content perceived as emotionally neutral might be more likely to be disseminated among peers.

Positive sentiment correlated strongly with supportive behaviors, showing significant associations with likes (*r* = 0.68) and coins (*r* = 0.74). This indicates that positively perceived content substantially encourages supportive interactions from users. Nevertheless, the impact of positive sentiment on view counts and comment volumes was comparatively weaker, suggesting that positive emotions mainly drive supportive engagement rather than general interaction or debate.

## Discussion

This study provides a data-driven exploration of adolescents’ reproductive health concerns and engagement on a major social media platform. The analysis of Bilibili videos and user comments revealed several noteworthy findings. First, user engagement varied significantly by topic. Less commonly discussed subjects, for instance sterilization surgery or FABMs, generated disproportionate levels of interaction (likes, shares, and comments) relative to their frequency. In contrast, widely covered topics such as condom use or general reproductive health education showed more modest engagement rates. This suggests that while prevalent topics dominate in volume, they do not necessarily elicit the strongest interest, indicating an unmet curiosity about niche or novel topics. Second, the sentiment analysis highlighted distinct emotional reactions to different reproductive health themes. Content on sensitive or controversial issues like induced abortion and new contraceptive methods elicited predominantly negative sentiments (each over 50% negative). Conversely, topics that invited personal empathy or were informational, for example videos about coping with spontaneous abortion or contraceptive education, saw higher proportions of positive sentiment. Third, an important structural finding was the low representation of medically trained uploaders. Only about 17% of uploaders in this adolescent reproductive health dataset had a confirmed medical background, raising concerns about the quality and accuracy of information being disseminated. This comprehensive overview of results underlines critical areas for discussion, particularly the gaps between adolescents’ information needs and the content currently available to them online.

Our findings reinforce and extend the growing body of evidence that social media has become a pivotal resource for adolescent reproductive health information worldwide. Consistent with our results on Bilibili, studies across diverse settings show that many young people now turn to online platforms for guidance on sensitive topics that may be inadequately addressed by traditional channels [20–22]. For example, an Indian survey of over 10,000 girls found that those with any social media exposure had significantly higher knowledge about sexual intercourse, contraception, and HIV/AIDS. Social media–using adolescents in that study had 1.3–2.2 times greater odds of accurate reproductive health knowledge compared to those not using these platforms [20]. Similarly, in Ethiopia, adolescents who engaged with social media were nearly twice as likely to utilize sexual and reproductive health services as their non-connected peers [21]. These patterns underscore the empowering potential of social media to fill information gaps and motivate health-seeking behavior, especially in contexts where formal education or services fall short. Indeed, the anonymity and accessibility of online media are often cited as major advantages, enabling youth to learn about puberty, contraception, or sex without fear of embarrassment. Social media’s influence is not universally positive, however—the literature also cautions that unmoderated exposure can introduce risks such as sexual misinformation or harmful content [22–25]. Our study adds nuance to this discourse by mapping how adolescents in China organically engage with a popular platform for reproductive health, offering a novel non-Western perspective to predominantly Western-focused research.

In line with global trends, the content landscape we observed on Bilibili was heavily skewed toward basic contraception and general sexual education, mirroring the emphasis seen in many youth health initiatives [26]. For instance, we found “condom” and general education tags accounted for over half of all video topics—a focus echoed in other social media environments. On TikTok, one analysis noted that roughly 15% of popular youth-oriented reproductive health videos dealt with birth control, a share only surpassed by relationship-themed content. This convergence suggests that fundamental topics like condom use and contraceptive advice consistently dominate online sexual health discourse, likely reflecting both high demand and the foundational nature of these subjects in prevention efforts. At the same time, our findings highlight notable content gaps. Important but more specialized topics—such as fertility awareness, female condoms, or spontaneous abortion—were comparatively underrepresented on Bilibili. This imbalance aligns with concerns raised in prior studies that adolescents often lack comprehensive knowledge on less commonly discussed methods and issues. In India, for example, only about 11% of girls had sufficient knowledge of modern contraceptive methods in one recent survey, hinting that topics beyond the basics remain inadequately addressed [20]. The dearth of content we observed on certain subjects therefore likely signals broader educational shortcomings. Notably, we found that when such under-covered topics were addressed, they tended to provoke strong engagement.

A key insight from our study is that videos on stigmatized or emotionally charged topics (like induced abortion) garnered disproportionately high engagement relative to their frequency. This finding resonates with observations from other contexts that adolescents actively seek out information on taboo subjects online when offline sources are inaccessible. Research from conservative or resource-limited settings has documented that youth often feel unable to obtain frank information about issues like abortion, sexual assault, or less common contraceptive options from parents, schools, or clinics [22]. In this vacuum, social media and online communities become crucial outlets where they can explore these topics anonymously and interact with peers. The heightened comment and share rates we observed for videos about induced abortion and sterilization suggest an unmet informational need being filled on Bilibili.

Our results thus support the notion that digital platforms can surface latent concerns that might otherwise remain unaddressed. They also highlight a potential opportunity: by identifying which topics elicit especially high interest (e.g., abortion), health educators and policymakers can target those areas for more robust online engagement or incorporate them into formal curricula to better meet adolescents’ needs. At the same time, the global literature urges caution—when young people must rely on peer-produced social media content for sensitive health information, the accuracy and quality of that information can vary widely. This makes it imperative for public health stakeholders to monitor and supplement these organic discussions with evidence-based resources.

Building on these findings, our study further explored adolescents’ emotional responses to various reproductive health topics and their relationship with engagement behaviors on social media. We observed significant variations in sentiment across topics, with content addressing sensitive or controversial issues, such as induced abortion and novel contraceptive methods, eliciting predominantly negative sentiments from viewers. More than half of the comments on these topics expressed anxiety, fear, or criticism, aligning with research from Kenya that highlights how controversial reproductive health discussions frequently elicit intense emotional responses, driven by societal taboos and personal ethical concerns [27]. In contrast, videos perceived as informative or emotionally supportive—such as those addressing coping with spontaneous abortion or providing general contraceptive education—generated a higher proportion of positive sentiments, affirming prior research highlighting adolescents’ preference for practical guidance and emotional reassurance.

Furthermore, our analysis revealed nuanced correlations between user sentiment and specific engagement metrics. Negative sentiments were moderately correlated with increased comment volumes, reinforcing that emotionally provocative content stimulates active discussion among adolescents. Prior social media analyses indicate that contentious, negatively toned content spurs more vigorous discussion. For example, researchers observed that posts with negative sentiment elicited significantly higher comment activity, as users engaged in debate and dialogue around the controversial content [28]. Positive sentiments, however, showed strong associations with supportive interactions like likes and coins, suggesting adolescents visibly endorse content they find reassuring or helpful. Notably, neutral sentiments correlated positively with sharing behaviors, implying adolescents favor disseminating unbiased or informational content, potentially to avoid controversy or personal stigma.These insights collectively underscore the importance of strategic emotional framing in digital reproductive health education to enhance adolescent engagement and facilitate informed discussions on sensitive health topics.

Another important aspect of our findings is the role of uploaders’ backgrounds in shaping the information ecosystem. We discovered that a majority of high-engagement reproductive health videos on Bilibili were produced by individuals without formal medical credentials. In some topic categories, over 80%–90% of content came from laypersons (e.g., vloggers, peer educators, or amateurs), with only a minority of videos attributed to healthcare professionals or organizations. This imbalance is broadly consistent with trends reported on other platforms. For instance, an analysis of contraceptive discussions on Twitter found that roughly half of tweets were authored by ordinary consumers, whereas only 6% came from official health or news sources [29]. Conversely, there are emerging examples where professional-led content has gained traction: on TikTok, one study observed that health providers were behind a substantial portion of popular contraception videos, which collectively garnered millions of views [30]. Importantly, those provider-created TikTok videos were rated significantly higher in accuracy and quality than videos made by non-professionals. This echoes a common theme in the literature—while adolescents appreciate relatable, authentic voices on social media, they also risk exposure to myths or inaccuracies when expertise is lacking. In our study’s context, the prominence of non-expert uploaders on Bilibili raises both concerns and possibilities. On one hand, the prevalence of lay content might contribute to the propagation of misinformation or incomplete knowledge [31]. On the other hand, the popularity of these uploaders suggests they have struck a chord with the target audience in ways traditional sources perhaps have not. Their approachable style, personal storytelling, or cultural relevance may be key to engaging Chinese Gen Z viewers. The challenge, therefore, is not to displace peer educators, but to integrate more evidence-based messaging into these social media streams. This could involve collaborations where medical professionals partner with popular influencers or provide them with verified information to share, a strategy that some public health groups are already testing. Indeed, the researchers argue that health authorities must proactively join these online conversations—for instance, by debunking contraception myths in comment threads or posting their own content—to ensure adolescents receive accurate guidance amid the noise [29]. Our findings reinforce this imperative, showing a clear gap between the high interest in reproductive health content and the relatively low visibility of authoritative sources.

Our study presents several limitations that should be acknowledged. First, the dataset was collected exclusively from Bilibili, a platform primarily used by young individuals in China. Thus, the findings may not generalize to adolescents who use other social media platforms or reside in different cultural contexts. Second, the study’s cross-sectional design limits our ability to infer causal relationships from observed associations between engagement metrics, sentiment responses, and video content.

Furthermore, although sentiment analysis was performed using an advanced LLM (Qwen2.5-32B), the accuracy might still be affected by informal, colloquial, or nuanced expressions commonly encountered in social media contexts.

Lastly, the dataset only includes videos published up until June 2024; therefore, any changes in user engagement or sentiment that occur after this time could not be captured. Ongoing monitoring and longitudinal studies would be beneficial to continuously assess and understand the evolving reproductive health information needs and perspectives of adolescents on social media platforms.

## Conclusion

This study highlights significant insights into adolescents’ reproductive health concerns and engagement behaviors on social media, particularly through the analysis of user-generated content on Bilibili. The findings underscore that user interactions significantly vary by reproductive health topics, with sensitive or less commonly discussed subjects eliciting disproportionately higher engagement. Notably, emotionally charged or controversial issues such as “induced abortion” and “novel contraceptive methods” generated predominantly negative sentiments, emphasizing societal taboos and ethical apprehensions among adolescents.

In contrast, content that provided informational support or emotional reassurance—such as coping strategies for spontaneous abortion or general contraceptive education—elicited higher positive responses and supportive interactions. Moreover, this study identified a critical gap in content credibility, with only a minority of uploaders having verified medical backgrounds, potentially compromising the quality and accuracy of reproductive health information disseminated on the platform.

Given the substantial influence of social media as a primary resource for adolescents’ reproductive health education, collaborative efforts among healthcare professionals, educators, and policymakers are essential. It is crucial to enhance the accuracy and reliability of online health information by promoting greater involvement of medically qualified contributors. Furthermore, strategic emotional framing in digital reproductive health content should be emphasized to facilitate positive adolescent engagement and informed discussions on sensitive topics.

## Data Availability

All data produced in the present study are available upon reasonable request to the authors

## Acknowledgments

This section is intended only for general acknowledgements and thanks. Any information related to funding, data availability, author contributions, etc. should be entered directly into their dedicated fields in the PLOS Editorial Manager submission system, which will then be incorporated into the appropriate section in your article during the production process.

## Supporting information

S1 Table. Keywords for video search on Bilibili.

